# Peak Nasal SARS-CoV-2 and Influenza Viral Loads Relative to Symptom Onset, 2022-2025: Impact of Vaccination and Implications for Multiplexed Testing

**DOI:** 10.64898/2025.12.23.25342938

**Authors:** Joanne H. Hunt, Gregory L. Damhorst, Richard Parsons, Jennifer K. Frediani, Kaleb B. McLendon, Julie Sullivan, Adrianna L. Westbrook, Wilbur A. Lam, Greg S. Martin, Nira R. Pollock

## Abstract

**Background:** We previously reported that nasal SARS-CoV-2 viral loads (VL) peaked around the fourth day of symptoms in highly immune adults sampled April 2022 – April 2023, while influenza A VL peaked soon after symptom onset. We hypothesized that SARS-CoV-2 kinetics may have changed due to reduced COVID-19 incidence and altered vaccination patterns. Understanding how viral kinetics evolve over time is essential to inform testing strategies.

**Methods:** Participants with symptomatic upper respiratory infection were recruited at testing centers in Georgia April 14, 2023-April 11, 2025. Participants reported date of symptom onset and vaccination history. A nasal swab was tested on the Xpert® Xpress CoV-2/Flu/RSV-Plus assay and Ct values recorded.

**Results:** 552 adults (≥16 years) and 60 children (<16y) were SARS-CoV-2-positive; lowest median Ct values (highest VL) were observed on the 2^nd^ symptomatic day. Adults vaccinated within 12M (n=106) had peak VL on the 3^rd^ day, and those vaccinated ≥12M prior (n=334), on the 2^nd^. In influenza A-positive adults (n=181) and children (n=147), median VL peaked on the 1^st^ symptomatic day, versus the 4^th^ day in 157 influenza B-positive participants.

**Conclusions:** In 2023-25, median SARS-CoV-2 VL peaked earlier relative to symptom onset compared to a highly immune 2022-23 cohort. More recent vaccination correlated with delayed peak VL, suggesting more robust immunity correlates with earlier symptom onset relative to peak. Median influenza A VL were highest at symptom onset, versus 4 days into symptoms for influenza B. These findings can inform use of multiplexed antigen tests amidst changing immunity and viral circulation patterns.

**Summary Point:** In 2023-25, median nasal SARS-CoV-2 viral loads peaked around the second day of symptoms, versus the fourth day in a 2022-23 highly immune cohort. Median influenza A viral loads peaked soon after symptom onset, versus on the fourth symptomatic day for influenza B. These observations have implications for multiplexed testing.

## Introduction

Early in the COVID-19 pandemic, multiple studies assessed viral kinetics relative to symptom onset and observed that SARS-CoV-2 viral loads (VLs) in upper respiratory samples, as approximated from polymerase chain reaction (PCR) cycle threshold (Ct) values, appeared to peak at symptom onset and decline steadily thereafter [1–4]. This observation informed early guidance on testing and contact tracing, with the two days prior to symptom onset considered a high-risk period for transmission. In contrast, studies conducted later in the pandemic after the acquisition of substantial natural and vaccine-derived immunity indicated that peak VL was later, occurring 3-4 days after the onset of COVID-19 symptoms [5–7]. These findings suggested that in a highly immune population, a rapid and robust immune response early in SARS-CoV-2 infection could lead to the early development of symptoms, with symptom onset thus preceding peak VL by a few days [8]. Cumulatively, these observations of delayed peak SARS-CoV-2 VL relative to symptom onset have had implications for testing practice [8]. This is of particular importance for the use of rapid antigen tests that are not sensitive enough to detect very low VLs and therefore may miss early infection [8]. Consistent with these observations, the current FDA recommendation is for symptomatic individuals to repeat rapid antigen testing 48h after an initial negative test [9].

In a previous study, we utilized data from symptomatic adults (defined as ≥16 years of age) presenting to hospital and community-based testing sites (April 2022-April 2023) to characterize the viral kinetics of SARS-CoV-2 and influenza A relative to symptom onset [5]. We evaluated SARS-CoV-2 VL distributions among PCR-positive adults by Ct value and antigen concentration in this highly exposed cohort, with over 90% having prior COVID-19 immunity (from vaccination, infection, or both) [5]. Median SARS-CoV-2 VLs in this cohort increased from those at symptom onset and peaked 4-5 days later. In influenza A PCR-positive adults, median influenza VLs peaked on the second day of symptoms. This analysis provided evidence to inform ongoing testing strategies for both COVID-19 and influenza A [5].

As we move further into a post-pandemic era, marked by lower documented SARS-CoV-2 infection rates, unpredictable waves of novel variants, and likely less frequent vaccination, understanding the viral kinetics of SARS-CoV-2 relative to symptom onset is essential to inform testing strategies and to maintain our ongoing understanding of COVID-19 transmission dynamics. In contrast, the annual timing of influenza circulation has remained largely stable, with relatively consistent influenza vaccination strategies and messaging. We thus took the opportunity to analyze data from the most recent cohort of symptomatic individuals tested for COVID-19, influenza A, and influenza B in the same testing program that contributed to our prior study to assess viral kinetics relative to symptom onset in a more recent population. In addition to this attempt to redefine the “new normal,” we expanded our analysis to characterize kinetics in individuals <16 years of age and to ask whether recent vaccination alters these patterns in a largely exposed population.

## Methods

The Atlanta Center for Microsystems Engineered Point-of-Care Technologies network utilizes hospital and community-based testing sites to enroll participants with symptomatic upper respiratory infection for evaluation of novel viral diagnostic tests under development (i.e. the “parent study”). This parent study has been running continuously since 2020 [10]. The parent study included any person seeking testing for an upper respiratory infection, and all individuals who were unable to tolerate a nasal swab or provide informed consent were excluded. Additional recruitment details are in Supplementary Methods. Demographic and clinical variables included age, sex, symptoms and date of symptom onset, location at time of enrollment, and vaccination status (including date of most recent SARS-CoV-2 and/or influenza vaccines).

For all participants in the parent study, an FDA-approved reverse transcription polymerase chain reaction (RT-PCR) SARS-CoV-2/influenza multiplexed test was performed within approximately 24 hours of study enrollment and nasal swab sample collection.

For the current analysis, existing data available from the parent study were retrospectively analyzed using a specific set of exclusion and inclusion criteria. Only data from participants tested with the Xpert Xpress SARS-CoV-2/Flu/RSV *plus* RT-PCR assay (Cepheid, Sunnyvale, CA) were included. Our prior analysis [5] of 2022-23 data had separately analyzed participants with and without known positive testing for COVID-19 in the 14 days prior to enrollment. For the current re-analysis of SARS-CoV-2 data from the same 2022-23 window, we included all participants from the parent study who had positive results of study PCR testing (non-zero Ct value) for SARS-CoV-2. For analysis of parent study data between April 14, 2023, and April 11, 2025, we included all participants who had positive results of study PCR testing for SARS-CoV-2, influenza A, or influenza B. The Xpert assay generates singular Ct values for SARS-CoV-2 and influenza B targets; for influenza A, we analyzed the A1 Ct value. For the 2023-25 cohort, we also analyzed data for participants of all ages, classifying those <16 years as children and those ≥16 years as adults. Exclusion criteria for this analysis included a missing date for symptom onset, a symptom duration greater than 6 days, and participants with co-detection of SARS-CoV-2 and influenza A or B. For the SARS-CoV-2 analysis, we also excluded participants with missing COVID-19 vaccination status data.

The sample collection date and the self-reported date of symptom onset were used to calculate the number of days of symptoms the participant had experienced at the time of sample collection (with the day of symptom onset designated as day 0). Participants were analyzed in groups of 0-6 days since symptom onset.

Ct values were plotted using box-and-whisker plots to represent distribution, median, and interquartile range (IQR). To estimate the hypothetical performance of Ag RDTs in this population, we also calculated the percentage of participants with a Ct value ≤25 or ≥30.

All analyses were performed in R (Boston, MA, v4.4.3), and GraphPad Prism (Boston, MA, v10.0.0).

## Results

The prior analysis [5] examined data from parent study participants enrolled between April 1, 2022, and April 13, 2023, and the current analysis reexamines the data from this window and also examines data for participants enrolled between April 14, 2023, and April 11, 2025. To contextualize these data collection windows, we plotted a timeline of SARS-CoV-2 hospitalizations and influenza A and B cases across Georgia reported to the COVID-19 Hospitalization Surveillance Network (COVID-NET) and Influenza Hospitalization Surveillance Network (FluSurv-NET) from January 2022 to April 2025 (**Figure 1**) [11].

**Figure 1.**
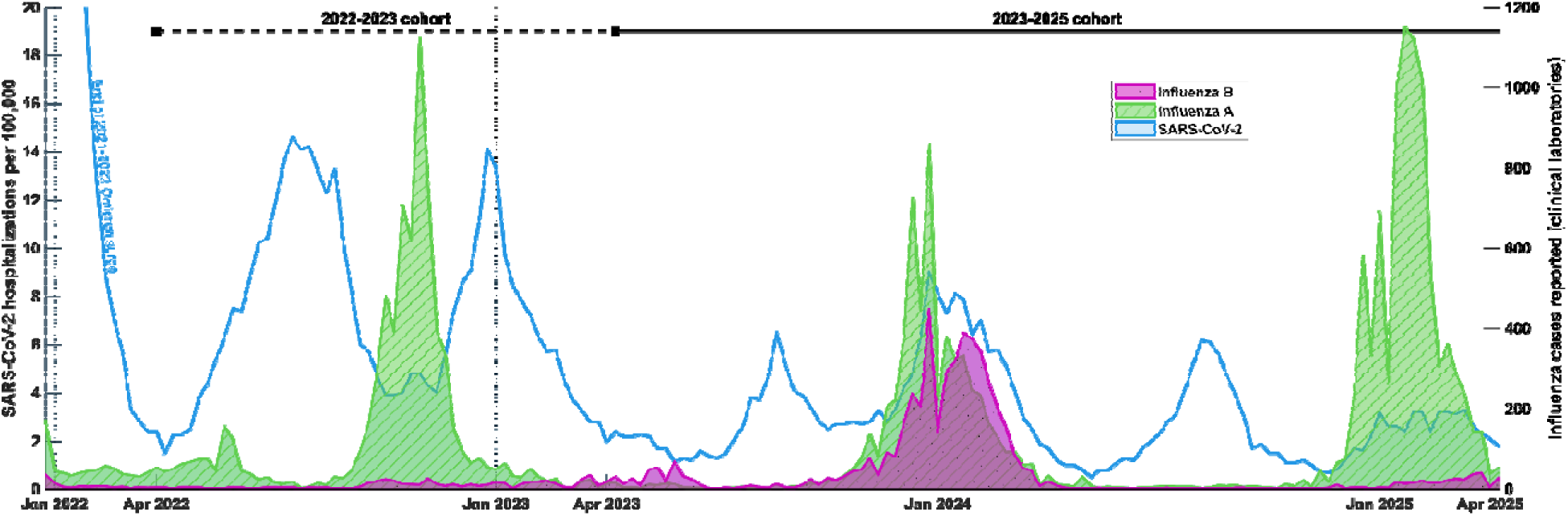
Plotted timeline of SARS-CoV-2 hospitalizations and influenza. A and B cases reported from clinical laboratories in Georgia, January 2022 to April 2025 (COVID-NET and FluSurv-NET Surveillance System). The dashed line represents the 2022-23 window of data collection (originally presented in Frediani et al., [5]) and the solid line represents the 2023-25 window of data collection.

### SARS-CoV-2

Our analysis included 595 adult participants who were PCR-positive for SARS-CoV-2 in the 2022-23 cohort and 552 such participants in the 2023-25 cohort (**Table 1**). Most participants had been tested on the second, third, or fourth day of symptoms. In the 2022-23 cohort, 68% of the participants were vaccinated within the previous 12 months. In the 2023-25 cohort, the timing of most recent vaccination varied, ranging from 19% within the previous 12 months, 32% within the previous 12-24 months, and 28% more than 2 years prior. Most participants in both cohorts were enrolled in community and urgent care settings (**Table 1**). Data for SARS-CoV-2 PCR-positive children in the 2023-25 cohort are presented in *Table S1* (notably, most did not have history of vaccination).

**Table 1.** Characteristics of SARS-CoV-2 PCR-Positive Adult Participants (≥16 years of age) in the 2022-2023 Cohort and the 2023-2025 Cohort.

| **Characteristic** | **2022-2023**<br>N = 595 | **2023-2025**<br>N = 552 |
| --- | --- | --- |
| Sex |  |  |
| Female | 380 (64) | 375 (68) |
| Male | 215 (36) | 177 (32) |
| Duration Since Symptom Onset, days <sup>a</sup> |  |  |
| 0 | 30 (5) | 31 (6) |
| 1 | 136 (23) | 137 (25) |
| 2 | 159 (27) | 181 (33) |
| 3 | 122 (21) | 119 (22) |
| 4 | 69 (12) | 55 (10) |
| 5 | 43 (7) | 21 (4) |
| 6 | 36 (6) | 8 (1) |
| Time since most recent COVID-19 Vaccination |  |  |
| Never vaccinated | 80 (13) | 94 (17) |
| Unknown duration | 23 (4) | 18 (3) |
| <12 months | 407 (68) | 106 (19) |
| 12-24 months | 82 (14) | 178 (32) |
| >24 months | 3 (<1) | 156 (28) |
| Testing Location |  |  |
| Community and Urgent Care Sites | 411 (69) | 301 (55) |
| Hospital-based Sites<sup>b</sup> | 184 (31) | 248 (45) |
| Other | 0 | 3 (<1) |

<sup>a</sup> Duration since symptom onset was determined as the difference between the sample collection date and the self-reported date of symptom onset (with the day of symptom onset designated as day 0).
<sup>b</sup> “Hospital-based sites” includes patients in the emergency department, ambulatory clinics located at hospital facilities, and admitted inpatients. The disposition (hospital admission or discharge home) was not recorded for emergency department patients.

Figure 2 shows SARS-CoV-2 Ct values from testing of nasal swab samples from symptomatic adult participants plotted by the duration of symptoms at the time the sample was collected. Using Ct value as a proxy measure for VL in the sample, lower Ct values correspond to higher nasal SARS-CoV-2 VL. The 2022-23 adult cohort demonstrated a peak median VL on day 4 of symptoms (Figure 2A), consistent with the prior analysis of these data [5]. In contrast, the 2023-25 adult cohort demonstrated a shift earlier with a peak median VL on day 2 of symptoms (Figure 2B); a similar trend was observed in children from this cohort (*Figure S1*).

**Figure 2.**
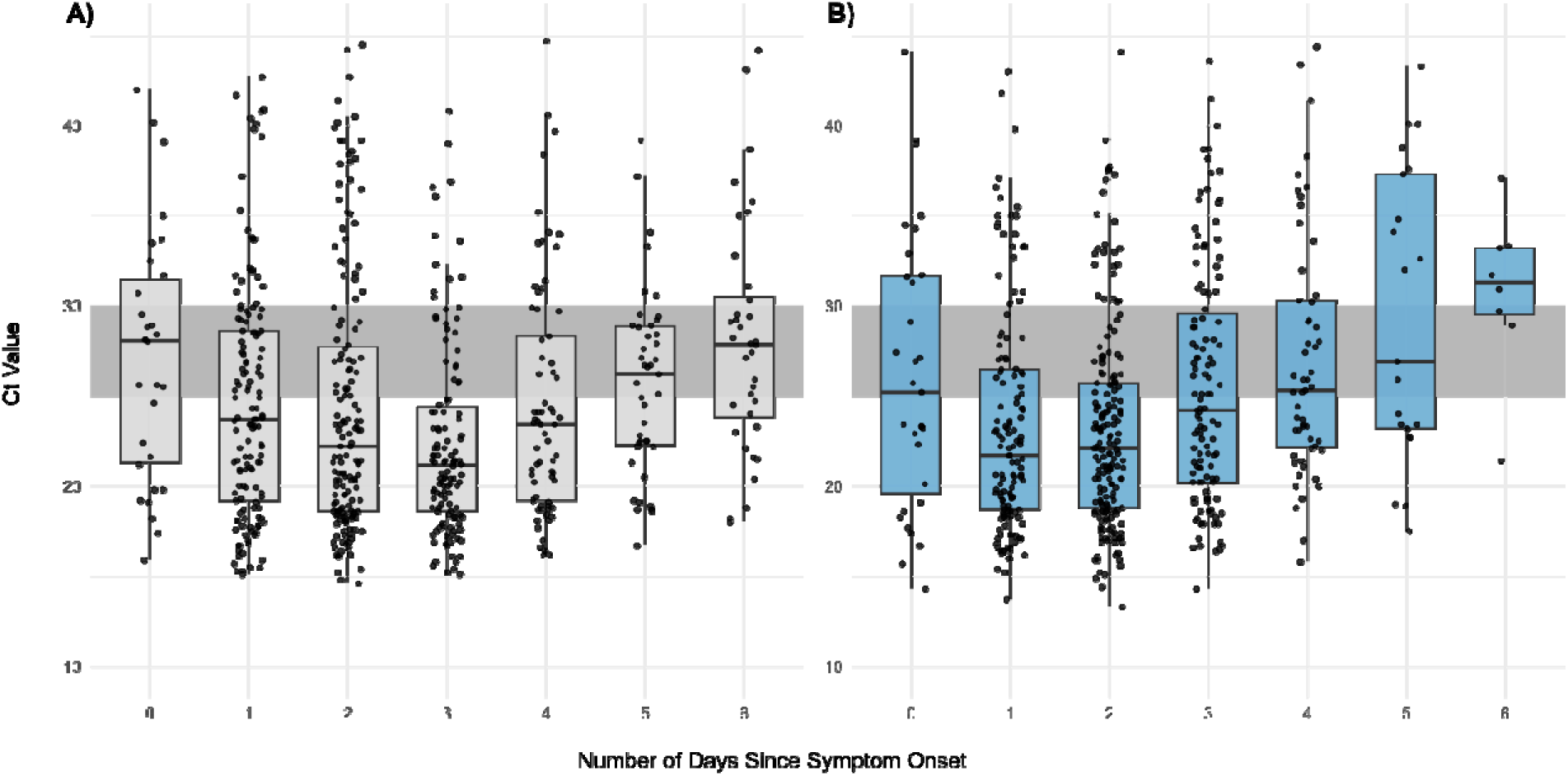
SARS-CoV-2 Ct values measured in nasal swab samples plotted by days since symptom onset for PCR-positive symptomatic adults (day 0 = the first day of symptoms). Panel A, 2022-23 cohort; Panel B, 2023-25 cohort. The grey bars in each panel outline the window between Ct of 30 and Ct of 25. *Abbreviations: Ct, Cycle threshold; PCR, polymerase chain reaction*.

To investigate the impact of vaccination timing within the 2023-25 adult cohort, we stratified by the time since most recent vaccination (Figure 3). Participants with most recent vaccination within the previous 12 months had peak median VL on the third day of symptoms (Figure 3A). For those with increasing time since vaccination, peak VL shifted earlier relative to symptom onset (day 2 of symptoms for those vaccinated between 12-24 months or >24 months prior; Figure 3B and 3C).

**Figure 3.**
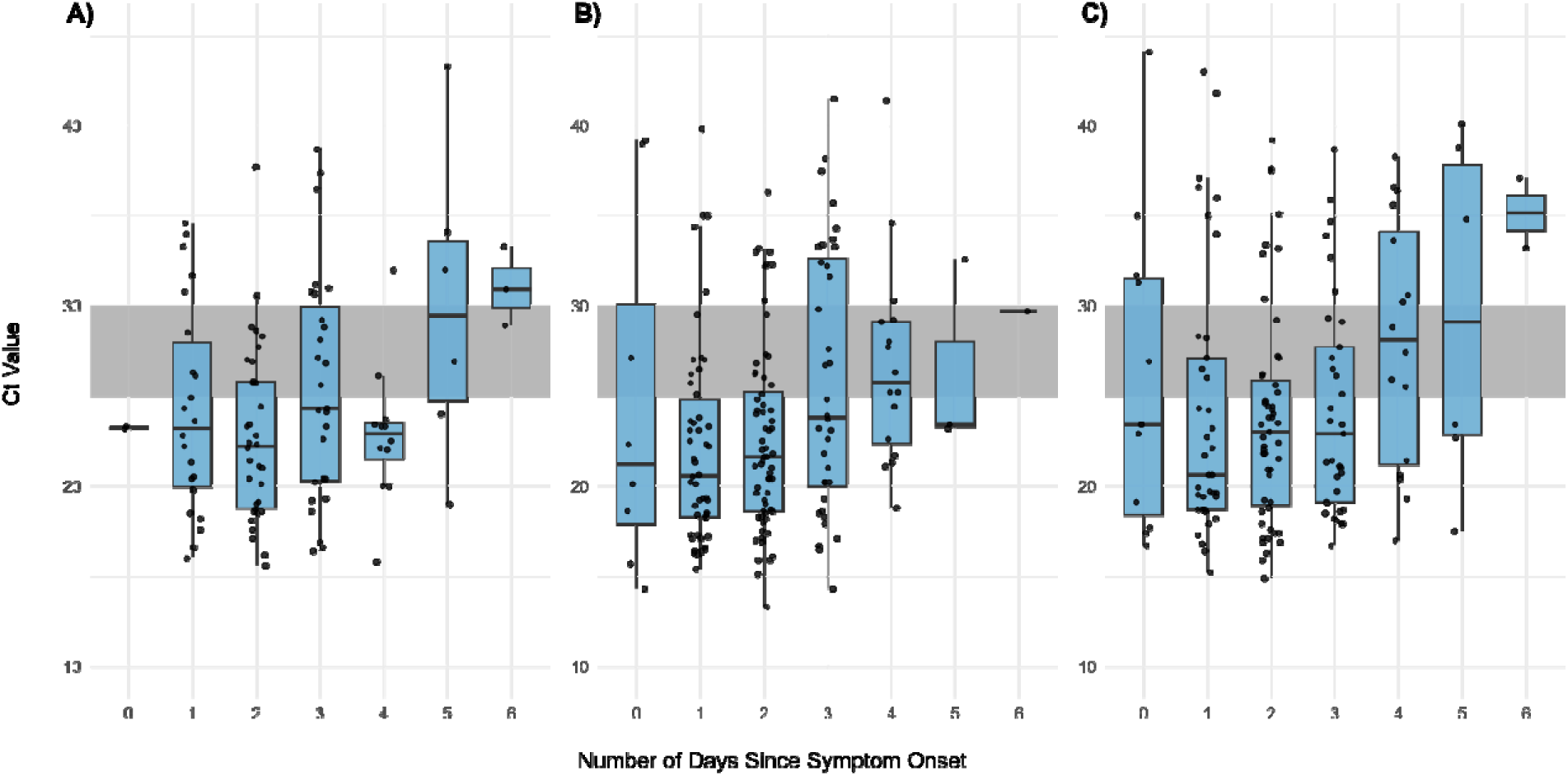
SARS-CoV-2 Ct values measured in nasal swab samples plotted by days since symptom onset for PCR-positive symptomatic adults (day 0 = the first day of symptoms) in the 2023-25 cohort, grouped by time since most recent vaccination. Time since last vaccination was determined as the difference between the self-reported most recent vaccination date and the date of sample collection. Panel A, less than 12 months since most recent vaccination (n=106); Panel B, 12-24 months since most recent vaccination (n=178); Panel C, over 24 months since most recent vaccination (n=156). The grey bars in each panel outline the window between Ct of 30 and Ct of 25. *Abbreviations: Ct, Cycle threshold; PCR, polymerase chain reaction*.

To estimate the percentage of PCR-positive adults expected to test positive on an antigen rapid diagnostic test (Ag RDT) on each day following symptom onset, we calculated the percentage of adults with a Ct value ≤30 and ≤25 for each day (**Table 2**). As done previously [5], we chose these thresholds based on internal data to represent the most sensitive (detecting samples with Ct ≤30) and least sensitive (detecting Ct ≤25) Ag RDTs available, though currently available Ag RDTs can occasionally detect samples with Ct >30. For the 2022-23 cohort, the highest estimated sensitivity of Ag RDT was predicted to be 77-90% on the fourth day of symptoms, while in the 2023-25 cohort, the highest sensitivity was predicted to be on the second (69-83%) or third (71-86%) day of symptoms. Similar results were observed in children from the 2023-25 cohort (*Table S2*), with highest predicted Ag RDT sensitivity on the second (75-88%) or third (68-89%) day of symptoms.

**Table 2.** SARS-CoV-2 Cycle Threshold (Ct) values, Number/Percent of Adult Samples (≥16 years of age) with Ct Values no more than Ct 25 or Ct 30, Grouped by Days Since Symptom Onset.

| Duration Since<br>Symptom Onset, days | 2022-2023 Cohort |  |  |  | 2023-2025 Cohort |  |  |  |
| --- | --- | --- | --- | --- | --- | --- | --- | --- |
| | n | Median (IQR) | $\leq 25$ Ct, n (%) | $\leq 30$ Ct, n (%) | n | Median (IQR) | $\leq 25$ Ct, n (%) | $\leq 30$ Ct, n (%) |
| 0 | 30 | 28.1 (21.3, 31.5) | 11 (37) | 21 (70) | 31 | 25.2 (19.6, 31.7) | 15 (48) | 21 (68) |
| 1 | 136 | 23.7 (19.2, 28.6) | 76 (56) | 113 (83) | 137 | 21.7 (18.7, 26.5) | 94 (69) | 114 (83) |
| 2 | 159 | 22.2 (18.6, 27.7) | 103 (65) | 126 (79) | 181 | 22.1 (18.8, 25.7) | 128 (71) | 156 (86) |
| 3 | 122 | 21.2 (18.6, 24.4) | 94 (77) | 110 (90) | 119 | 24.2 (20.2, 29.6) | 63 (53) | 90 (76) |
| 4 | 69 | 23.4 (19.2, 28.3) | 45 (65) | 55 (80) | 55 | 25.3 (22.2, 30.3) | 25 (45) | 40 (73) |
| 5 | 43 | 26.2 (22.3-28.9) | 19 (44) | 37 (86) | 21 | 26.9 (23.2, 37.3) | 9 (43) | 11 (52) |
| 6 | 36 | 27.9 (23.8, 30.5) | 12 (33) | 26 (72) | 8 | 31.3 (29.5, 33.2) | 1 (13) | 3 (38) |

Abbreviations: IQR, Interquartile range; Ct, Cycle threshold.

### Influenza A and B

There were 147 children and 181 adults who were PCR-positive for influenza A in the 2023-25 cohort (**Table 3**). Most children (73%) and adults (65%) had not received influenza vaccination within the previous 12 months. Most participants were tested on the second, third, or fourth day of symptoms. More than half were enrolled at hospital-based sites.

**Table 3.** Characteristics of Influenza A PCR-Positive Participants in the 2023-2025 Cohort.

| Characteristic | <16 Years of Age<br>N = 147 | ≥16 years of Age<br>N = 181 |
| --- | --- | --- |
| Sex |  |  |
| Female | 68 (47) | 98 (54) |
| Male | 78 (53) | 82 (45) |
| Unknown |  | 1 (<1) |
| Duration Since Symptom Onset, days<sup>a</sup> |  |  |
| 0 | 7 (5) | 10 (6) |
| 1 | 35 (24) | 38 (21) |
| 2 | 32 (22) | 61 (34) |
| 3 | 29 (20) | 46 (25) |
| 4 | 24 (16) | 16 (9) |
| 5 | 17 (12) | 7 (4) |
| 6 | 3 (2) | 3 (2) |
| Flu Vaccination in Past 12 months |  |  |
| Yes | 33 (22) | 52 (29) |
| No | 107 (73) | 118 (65) |
| Unknown | 7 (5) | 11 (6) |
| Testing Location |  |  |
| Community and Urgent Care Sites | 57 (39) | 71 (39) |
| Hospital-based Sites<sup>b</sup> | 90 (61) | 110 (61) |

<sup>a</sup>Duration since symptom onset was determined as the difference between the sample collection date and the self-reported date of symptom onset (with the day of symptom onset designated as day 0).
<sup>b</sup>“Hospital-based sites” includes patients in the emergency department, ambulatory clinics located at hospital facilities, and admitted inpatients. The disposition (hospital admission or discharge home) was not recorded for emergency department patients.

Figure 4 shows influenza A Ct values plotted by duration of symptoms, again using Ct value as a proxy for VL. In both children and adults, the peak median influenza A VL was on the first day of symptoms (**Figure 4A and 4B**). Similarly, we previously observed that median VL peaked on the second day of symptoms in adults [5].

**Figure 4.**
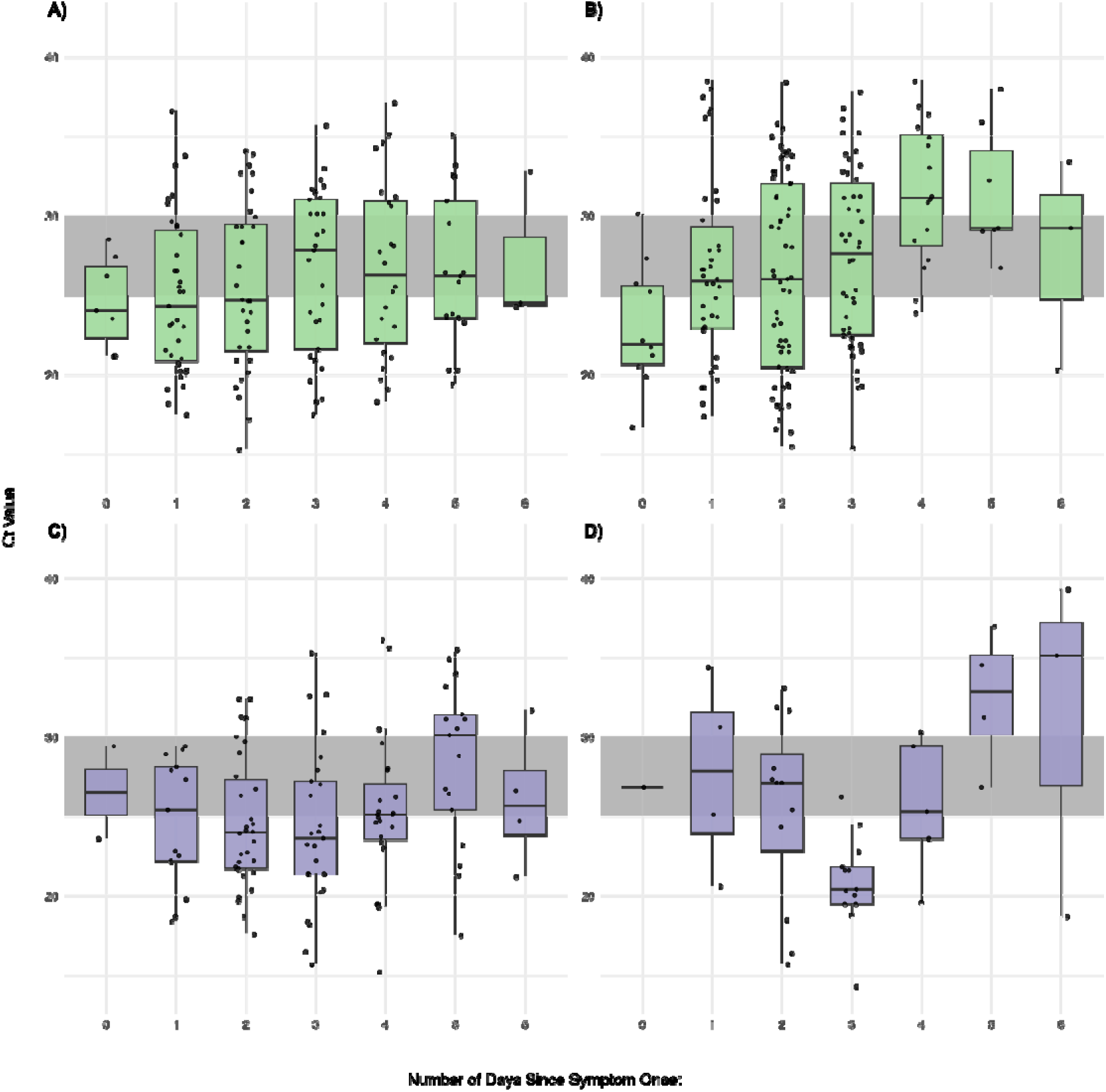
Influenza A and influenza B Ct values measured in nasal swab samples plotted by days since symptom onset for PCR-positive symptomatic participants (day 0 = the first day of symptoms) in the 2023-25 cohort. Panel A, influenza A-positive participants under the age of 16 years; Panel B, influenza A-positive participants 16 years and older; Panel C, influenza B-positive participants under the age of 16 years; Panel D, influenza B-positive participants 16 years and older. The grey bars in each panel outline the window between Ct of 30 and Ct of 25. *Abbreviations: Ct, Cycle threshold; PCR, polymerase chain reaction*.

While influenza vaccination in the prior 12 months did appear to have a potential correlation with a delayed peak influenza A VL (*Figure S2B*) compared to no vaccination in the prior 12 months (*Figure S2A*), sample size limited analysis.

In both children and adults, the highest predicted sensitivity for influenza A Ag RDT was on the day of symptom onset (day 0), ranging from 57-100% in children and 60-90% in adults (**Table 4**).

**Table 4.** Influenza A Cycle Threshold (Ct) Values, Number/Percent of Samples in the 2023-2025 Cohort with Ct Values no more than Ct 25 or Ct 30, Grouped by Age Stratification (<16 years and ≥16 Years) and Number of Days Since Symptom Onset.

| Duration Since Symptom<br>Onset, days | Children (<16 Years) |  |  |  | Adults (≥16 years) |  |  |  |
| --- | --- | --- | --- | --- | --- | --- | --- | --- |
|  | n (%) | Median (IQR) | ≤25 Ct, n (%) | ≤30 Ct, n (%) | n (%) | Median (IQR) | ≤25 Ct, n (%) | ≤30 Ct, n (%) |
| 0 | 7 | 24.0 (22.4, 26.8) | 4 (57) | 7 (100) | 10 | 21.9 (20.7, 25.6) | 6 (60) | 9 (90) |
| 1 | 35 | 24.3 (20.9, 29.1) | 18 (51) | 29 (83) | 38 | 25.9 (22.9, 29.3) | 16 (42) | 29 (76) |
| 2 | 32 | 24.7 (21.5, 29.5) | 17 (53) | 25 (78) | 61 | 26.0 (20.5, 32.0) | 27 (44) | 41 (67) |
| 3 | 29 | 27.8 (21.6, 31.0) | 12 (41) | 18 (62) | 46 | 27.6 (22.5, 32.0) | 19 (41) | 29 (63) |
| 4 | 24 | 26.3 (22.0, 30.9) | 10 (42) | 16 (67) | 16 | 31.1 (28.1, 35.1) | 2 (13) | 6 (38) |
| 5 | 17 | 26.2 (23.6, 30.9) | 7 (41) | 12 (71) | 7 | 29.2 (29.1, 34.1) | 0 | 4 (57) |
| 6 | 3 | 24.5 (24.4, 28.7) | 2 (67) | 2 (67) | 3 | 29.2 (24.8, 31.3) | 1 (33) | 2 (67) |

There were 115 children and 42 adults who were PCR-positive for influenza B (**Table 5**); 44% of children and 48% of adults reported no influenza vaccination within the previous 12 months. Most children were tested on the third, fourth, or fifth day of symptoms and most adults on the third or fourth day. More than half were enrolled at hospital-based sites.

**Table 5.**
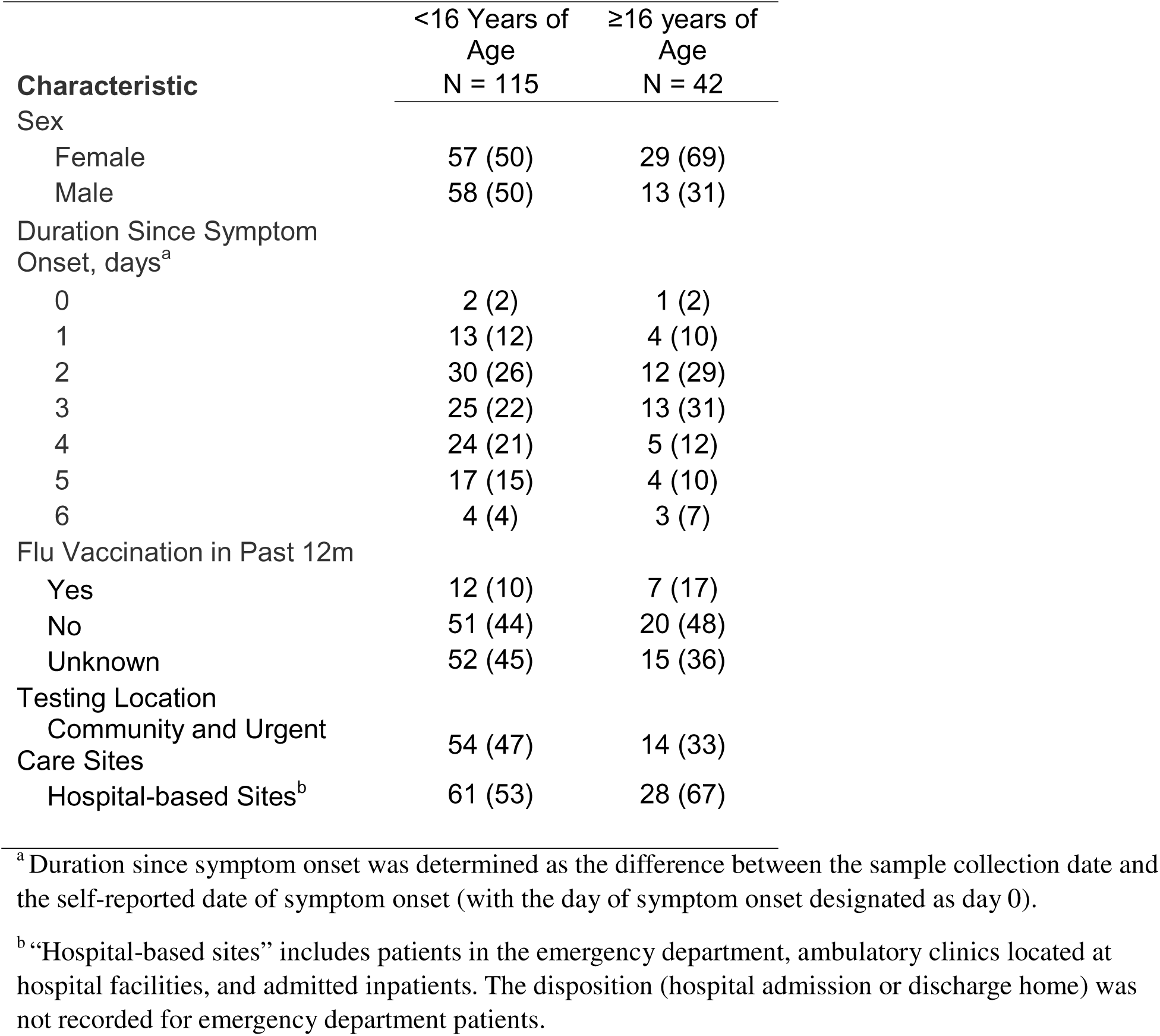
Characteristics of Influenza B PCR-Positive Participants in the 2023-2025 Cohort.

| Characteristic | <16 Years of Age<br>N = 115 | ≥16 years of Age<br>N = 42 |
| --- | --- | --- |
| Sex |  |  |
| Female | 57 (50) | 29 (69) |
| Male | 58 (50) | 13 (31) |
| Duration Since Symptom Onset, days<sup>a</sup> |  |  |
| 0 | 2 (2) | 1 (2) |
| 1 | 13 (12) | 4 (10) |
| 2 | 30 (26) | 12 (29) |
| 3 | 25 (22) | 13 (31) |
| 4 | 24 (21) | 5 (12) |
| 5 | 17 (15) | 4 (10) |
| 6 | 4 (4) | 3 (7) |
| Flu Vaccination in Past 12m |  |  |
| Yes | 12 (10) | 7 (17) |
| No | 51 (44) | 20 (48) |
| Unknown | 52 (45) | 15 (36) |
| Testing Location |  |  |
| Community and Urgent Care Sites | 54 (47) | 14 (33) |
| Hospital-based Sites<sup>b</sup> | 61 (53) | 28 (67) |

<sup>a</sup> Duration since symptom onset was determined as the difference between the sample collection date and the self-reported date of symptom onset (with the day of symptom onset designated as day 0).
<sup>b</sup> “Hospital-based sites” includes patients in the emergency department, ambulatory clinics located at hospital facilities, and admitted inpatients. The disposition (hospital admission or discharge home) was not recorded for emergency department patients.

In contrast to influenza A, the influenza B peak median VL was observed on the 4^th^ day of symptoms in both children and adults (**Figure 4C and 4D**). Small sample size limited vaccination impact analysis (*Figure S2C/S2D*).

The highest predicted sensitivity of influenza B Ag RDT was on days 3 and 4 of symptoms for children and day 4 of symptoms for adults, though sample size limited analysis and conclusions (**Table 6**).

**Table 6.** Influenza B Cycle Threshold (Ct) Values, Number/Percent of Samples in the 2023-2025 Cohort with Ct Values no more than Ct 25 or Ct 30, Grouped by Age Stratification (<16 years and ≥16 Years) and Number of Days Since Symptom Onset.

| Duration Since Symptom Onset, days | Children (<16 Years) |  |  |  | Adults (≥16 years) |  |  |  |
| --- | --- | --- | --- | --- | --- | --- | --- | --- |
|  | n (%) | Median (IQR) | ≤25 Ct, n (%) | ≤30 Ct, n (%) | n (%) | Median (IQR) | ≤25 Ct, n (%) | ≤30 Ct, n (%) |
| 0 | 2 | 26.5 (25.1, 28.0) | 1 (50) | 2 (100) | 1 | 26.8 | 0 | 1 (100) |
| 1 | 13 | 25.4 (22.2, 28.1) | 6 (46) | 13 (100) | 4 | 27.9 (24.0, 31.6) | 1 (25) | 2 (50) |
| 2 | 30 | 24.0 (21.7, 27.3) | 20 (67) | 26 (87) | 12 | 27.1 (22.9, 28.9) | 4 (33) | 9 (75) |
| 3 | 25 | 23.6 (21.4, 27.2) | 16 (64) | 21 (84) | 13 | 20.4 (19.5, 21.8) | 12 (92) | 13 (100) |
| 4 | 24 | 25.1 (23.6, 27.1) | 11 (48) | 20 (87) | 5 | 25.3 (23.6, 29.4) | 2 (40) | 4 (80) |
| 5 | 17 | 30.1 (25.4, 31.4) | 4 (24) | 8 (47) | 4 | 32.9 (30.1, 35.1) | 0 | 1 (25) |
| 6 | 4 | 25.7 (23.8, 27.9) | 2 (50) | 3 (75) | 3 | 35.1 (26.9, 37.2) | 1 (33) | 1 (33) |

## Discussion

The existence of a long-running, consistently operated parent study assessing novel rapid diagnostic tests for respiratory viruses—first those for SARS-CoV-2, and then those multiplexed to also detect influenza A and B—has provided a unique opportunity to assess how the relationships between SARS-CoV-2 and influenza VL kinetics and the onset of symptoms have evolved over the past four years. The timing of peak nasal VL relative to symptom onset directly impacts test use, particularly for tests that are not sensitive enough to detect the low VLs present early in the course of infection. Accordingly, repeat testing (48h later) is currently required in a newly symptomatic patient who initially tests negative on a rapid COVID antigen test [9], and multiplexed SARS-CoV-2/influenza antigen tests have maintained serial testing requirements and/or suggest follow-up with a molecular test [12–14]. This analysis builds on our prior analysis of SARS-CoV-2 and influenza A VL in a 2022-23 adult cohort [5] and contributes new evidence from a 2023-25 cohort to inform current testing strategies for SARS-CoV-2, influenza A, and influenza B in adults and children.

In our prior analysis [5], we observed that nasal SARS-CoV-2 VL peaked around the fourth day of symptoms, while influenza A VL peaked soon after symptom onset. The observed SARS-CoV-2 kinetics were markedly different than those observed early in the COVID-19 pandemic, when VL in upper respiratory samples appeared to peak at symptom onset and then decline steadily [1–4]. In the 2022-23 cohort, 91% had history of COVID-19 vaccination, prior SARS-CoV-2 infection, or both [5]. At that time, given frequent COVID-19 vaccine boosters and high disease prevalence, any vaccination or prior infection was relatively recent. These findings and those of others [5–7] suggested that in a highly immune population, a rapid and robust immune response early in SARS-CoV-2 infection could lead to early symptom development that precedes peak VL by multiple days [8].

This analysis presents data from samples collected in a more recent population, allowing us to comparatively illustrate SARS-CoV-2 kinetics in a population where vaccination and natural infection are less common. We observed that median SARS-CoV-2 VL peaked earlier in the 2023-25 cohort, with peak VL occurring on the second day of symptoms rather than on the fourth. While we do not have data on history of prior infection in the 2023-25 cohort, we were able to assess the impact of vaccination timing on the relationship between symptom onset and peak VL. Notably, in the 2023-25 cohort, COVID-19 vaccination within the previous 12 months correlated with delayed peak VL. Taken together, our data continue to suggest that more robust immunity to SARS-CoV-2 correlates with earlier symptom onset relative to peak VL. This could mean that individuals with more robust immunity might take longer to test positive on an Ag RDT after symptom onset, and conversely, individuals without robust immunity might test positive sooner. While limited by sample size, overall trends in children with SARS-CoV-2 infection looked similar to adults.

Unlike the substantial changes in COVID-19 incidence and vaccination patterns over the past four years, the influenza A/B experience has remained relatively unchanged, with a late summer/early fall vaccine offering and mid-late fall onset of influenza A transmission in Georgia (though influenza B timing is more variable). We were able to expand our prior analysis to examine influenza A and B kinetics relative to symptom onset in both adults and children in 2023-25. Consistent with our findings from 2022-23 [5], adults positive for influenza A in 2023-25 had peak median VL on the first day of symptoms, indicating that testing early in the symptom course should have higher yield; the same trend was observed in children. These findings are consistent with a community-based study in Hong Kong (Ip et al, [15]), a household-based study in Wisconsin and Tennessee (Morris et al, [16]), and multiple human challenge studies [17] in which influenza A shedding peaked on the first 1-2 days of clinical illness. Most influenza A-positive participants reported no influenza vaccination within the previous 12 months. Although we observed some evidence of delayed peak VL with recent vaccination, small sample size limited analysis.

Unlike influenza A, for influenza B the peak median VL occurred on the fourth day of symptoms in both children and adults. Interestingly, Ip et al [15] noted a bimodal peak for influenza B VL, with one peak up to 2 days prior to symptom onset and a second around the 4^th^ or 5^th^ day of symptoms. Our observation of a delayed peak VL has significant implication for diagnosis of influenza B with rapid antigen tests, suggesting that such tests may not reliably detect influenza B until well into the symptom course.

Our analysis of the percentage of participants likely to test positive on Ag RDTs suggested that in the 2023-25 cohort, Ag RDTs for SARS-CoV-2 would have highest sensitivity on the second or third day of symptoms; influenza A RDTs would have highest sensitivity on the day of symptom onset; and influenza B RDTs would have highest sensitivity on day 3 of symptoms for children and day 4 for adults. Given that Ag RDTs detecting all three viruses simultaneously are now commercially available, our findings illustrate that the negative predictive value for each virus differs depending on when the test is performed relative to symptom onset.

Our study has some limitations. First, parent study data on prior positive testing were not consistently collected in 2023-25, limiting our ability to assess the impact of prior SARS-CoV-2 infection on VL kinetics in this analysis. Second, our study relied on self-reported symptom timing, which carries some degree of imprecision. Finally, for those enrolled in a hospital setting (ED, hospital-based clinic, or inpatient), the location within the hospital facility at the time of enrollment was not consistently documented, making it impossible to precisely determine the percentage of admitted patients.

Our data provide a starting point for redefinition of the “new normal” kinetics of SARS-CoV-2, influenza A, and influenza B in adults and children presenting with symptomatic upper respiratory infection. Additional studies evaluating the relationship between the timing of symptom onset and nasal viral load will be useful to confirm the kinetics observed. In the meantime, our results suggest that for multiplexed Ag RDTs detecting SARS-CoV-2, influenza A, and influenza B, serial testing (repeating testing 48h after an initial negative test in a patient with continued symptoms) and/or consideration of molecular testing in the right clinical context remain important.

## Data Availability

All data produced in the present work are contained in the manuscript.

## Contributors

NRP, GLD, and JHH designed the study. KBM performed and supervised laboratory testing. JKF, RP, JS, ALW, WAL and GSM supervised data collection and management. JHH performed data analyses. JHH, GLD, and NRP interpreted results. JHH, GLD, and NRP drafted the manuscript. NRP, JHH, GLD, JKF, and JS performed critical editing of the manuscript. All authors reviewed and approved the manuscript.

## Ethics Committee Approval

The study was approved by Emory University and Children’s Healthcare of Atlanta Institutional Review Boards.

