## Supplement File for "Peak Nasal SARS-CoV-2 and Influenza Viral Loads Relative to Symptom Onset, 2022-2025: Impact of Vaccination and Implications for Multiplexed Testing"

**Supplementary Methods**

Recruitment:

Given that one goal of the parent study was to assess the utility of novel test devices, patients with known SARS-CoV-2 or influenza infection were initially not actively recruited. As the study continued into a period of lower SARS-CoV-2 infection rates and consistent evaluation of tests detecting both SARS-CoV-2 and influenza viruses, the recruitment strategy was modified to enroll patients with known SARS-CoV-2 or influenza infection. Eligible participants were identified and contacted by study personnel who obtained informed consent.

Sample collection and testing:

Nasal swab samples for PCR testing were collected using a flocked swab (Copan 520CS01 or Puritan PurFlock Ultra 25-3606-U) in saline.

**Table S1.** Characteristics of SARS-CoV-2 PCR-positive children (<16 years of age) from the 2023-25 cohort.

| **Characteristic** | <16 Years (N = 60) |
| --- | --- |
|  | n (%) |
| Sex |  |
| Female | 35 (58) |
| Male | 25 (42) |
| Duration Since Symptom Onset, days ^a^ |  |
| 0 | 8 (13) |
| 1 | 16 (27) |
| 2 | 19 (32) |
| 3 | 9 (15) |
| 4 | 5 (8) |
| 5 | 2 (3) |
| 6 | 1 (2) |
| Time since most recent COVID-19 Vaccination |  |
| Never vaccinated | 35 (58) |
| Unknown duration | 3 (5) |
| <12 months | 4 (7) |
| 12-24 months | 6 (10 |
| >24 months | 12 (20) |
| Testing Location ^b^ |  |
| Community and Urgent Care Sites | 28 (47) |
| Hospital-based Sites | 32 (53) |

^a^ Duration since symptom onset was determined as the difference between the sample collection date and the self-reported date of symptom onset (with the day of symptom onset designated as day 0).

^b^ “Hospital-based sites” includes patients in the emergency department, ambulatory clinics located at hospital facilities, and admitted inpatients. The disposition (hospital admission or discharge home) was not recorded for emergency department patients.

**Table S2.** SARS-CoV-2 Cycle Threshold (Ct) Values in Children (<16 years of age), Number/Percent of Samples with Ct Values No More Than Ct 25 or Ct 30, Grouped by Number of Days Since Symptom Onset.

| Duration Since Symptom Onset, days | n | Median (IQR) | ≤25 Ct, n (%) | ≤30 Ct, n (%) |
| --- | --- | --- | --- | --- |
| 0 | 8 | 24.1 (21.0, 26.4) | 4 (50) | 8 (100) |
| 1 | 16 | 19.4 (18.6, 23.8) | 12 (75) | 14 (88) |
| 2 | 19 | 24.0 (19.7, 25.3) | 13 (68) | 17 (89) |
| 3 | 9 | 27.1 (23.1, 30.5) | 3 (33) | 6 (67) |
| 4 | 5 | 20.4 (19.5, 26.9) | 3 (60) | 4 (80) |
| 5 | 2 | 25.2 (24.1, 26.2) | 1 (50) | 2 (100) |
| 6 | 1 | 39.9 | 0 | 0 |

**Figure S1.** SARS-CoV-2 Ct values measured in nasal swab samples plotted by days since symptom onset for PCR-positive symptomatic children (<16 years) (day 0 = the first day of symptoms). The grey bars in each panel outline the window between Ct of 30 and Ct of 25. *Abbreviations: Ct, Cycle threshold: PCR, polymerase chain reaction.*

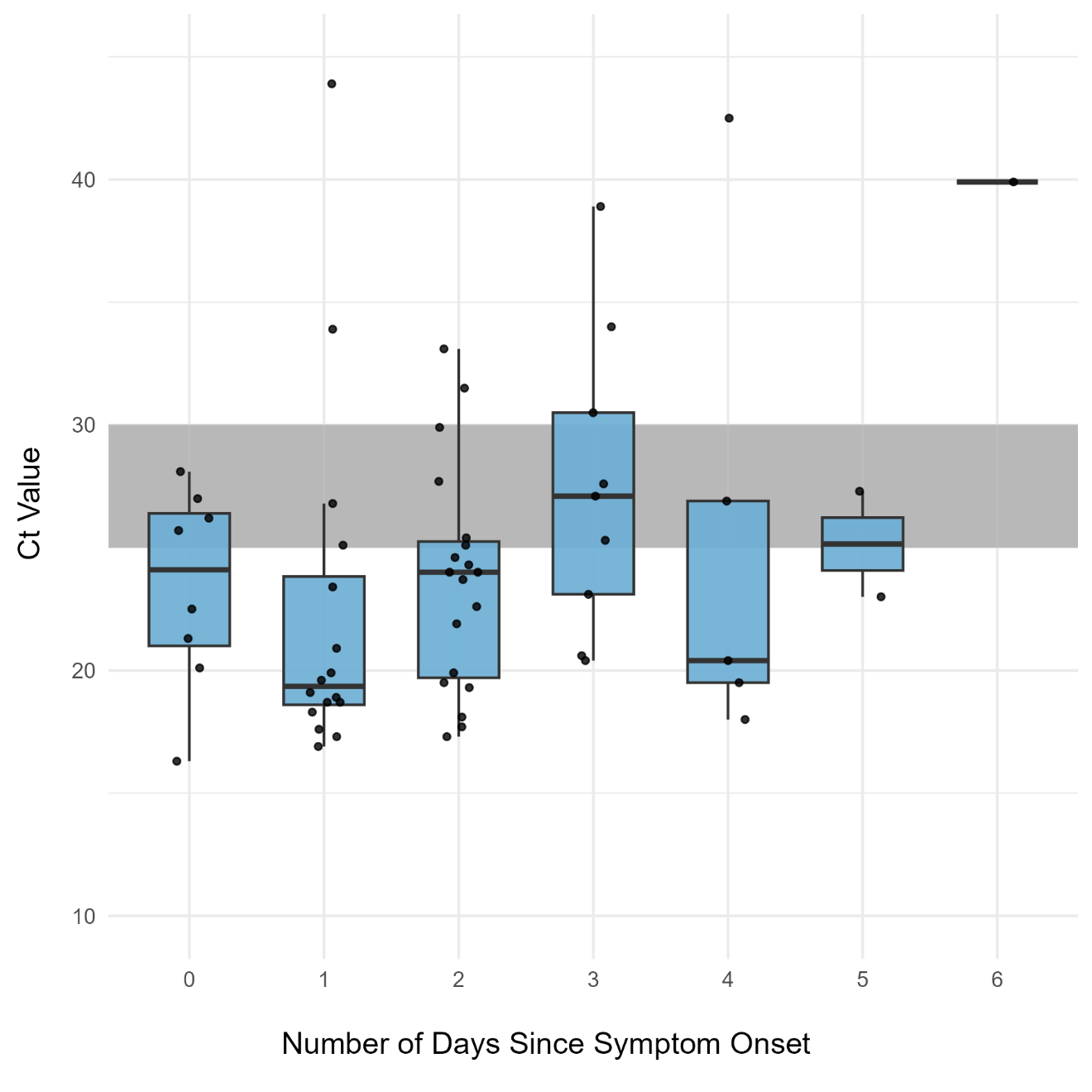

**Figure S2.** 2023-2025 Cohort Influenza A and B Ct values measured in nasal swab samples plotted by days since symptom onset for PCR-positive symptomatic adults and children (day 0 = the first day of symptoms), and by influenza vaccination in the last 12 months. Influenza A: Panel A (No influenza vaccination in previous 12 months) and Panel B (influenza vaccination in previous 12 months). Influenza B: Panel C (No influenza vaccination in previous 12 months) and Panel D (influenza vaccination in previous 12 months). The grey bars in each panel outline the window between Ct of 30 and Ct of 25. *Abbreviations: Ct, Cycle threshold: PCR, polymerase chain reaction.*

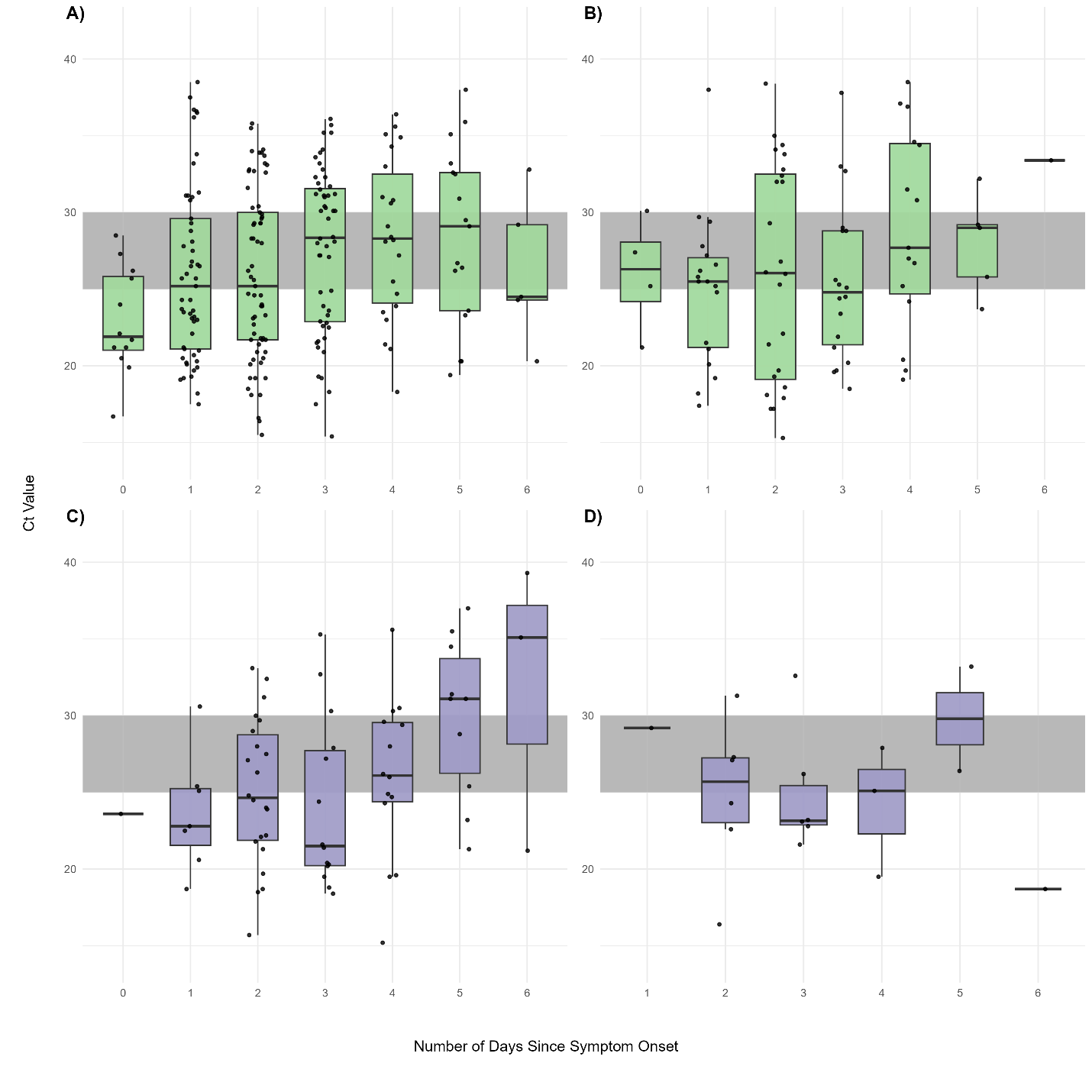
